# Epigenetic and cognitive signatures of striatal DAT binding among healthy elderly individuals

**DOI:** 10.1101/2024.10.15.24315507

**Authors:** Arash Yaghoobi, Homa Seyedmirzaei, Marzie Jamaat, Moein Ala

## Abstract

**Background:** Striatal dopamine transporter (DAT) binding is a sensitive and specific endophenotype for detecting dopaminergic deficits across Parkinson’s disease (PD) spectrum. Molecular and clinical signatures of PD in asymptomatic phases help understand the earliest pathophysiological mechanisms underlying the disease. We aimed to investigate whether blood epigenetic markers are associated with inter-individual variation of striatal DAT binding among healthy elderly individuals. We also investigated whether this potential inter-individual variation can manifest as dysfunction of particular cognitive domains.

**Method:** We conducted a blood epigenome-wide association study of striatal DAT binding on 96 healthy individuals using the Illumina EPIC array. For functional annotation of our top results, we employed the enhancer-gene mapping strategy using a midbrain single-nucleus multimodal dataset. Finally, we conducted several investigative regression analyses on several neuropsychological tests across five cognitive domains to assess their association with striatal DAT binding among 250 healthy subjects.

**Results:** We identified seven suggestive (P-value<10^−5^) CpG probes. Specifically, three probes were colocalized with three risk loci previously identified in PD’s largest Genome-Wide Association Study (GWAS). *UCN5A* and *APOE* loci were identified as suggestive DMRs associated with striatal DAT binding. Functional analyses prioritized the *FDFT1* gene as the potential target gene in the previously reported *CTSB* GWAS locus. We also showed that delayed recall memory impairment was correlated with reduced striatal DAT binding, irrespective of age.

**Conclusion:** Our study suggested epigenetic and cognitive signatures of striatal DAT binding among healthy individuals, providing valuable insights for future experimental and clinical studies of early PD.

## 1. Introduction

Dopamine transporter scan (DaTscan) is a nuclear imaging modality used to detect nigrostriatal dopaminergic deficits in Parkinson’s disease (PD) and other Parkinsonian syndromes [1]. PD is the second most common neurodegenerative disorder, affecting approximately 1% of people aged more than 60 years worldwide [2, 3]. It is estimated that the first motor symptoms of PD do not appear until approximately 35%-45% of striatal DAT activity is lost [4]. Therefore, PD is now seen as a continuum from asymptomatic individuals who are at high risk for PD to individuals with prodromal symptoms, all the way to PD patients. Considering this fundamental nature of PD, sensitive and specific biomarkers are valuable endophenotypes for quantifying the early stages of the disease [5]. Due to the high sensitivity (84.4%) and specificity (96.2%) of DaTscan [6], this imaging tool can serve as a robust endophenotype to quantify the amount of dopaminergic neuron loss in asymptomatic and prodromal phases of PD.

PD originates from pathological changes in various cells in the brain. These changes may be detected at the epigenomic level in the early stages of the disease [7]. One of the epigenetic mechanisms is DNA methylation, which controls gene expression. To date, few studies have proposed several genes whose methylations were altered in blood or brain tissues, such as *SNCA*, *DRD2*, *NPAS2*, *CRY1*, *MAPT*, and *PDE4D* [7]. However, these studies mostly compared the epigenomic profiles of patients with late-stage PD with healthy controls, which impedes our understanding of the biological pathways involved in early PD.

In addition to molecular changes in the brain, prodromal PD can manifest with clinical presentations, such as changes in cognition. It is proposed that approximately 40% of patients with early-stage PD suffer from mild cognitive impairment. A large longitudinal study on healthy subjects of the Rotterdam cohort has shown that cognitive dysfunction can increase the risk of future Parkinsonism among healthy individuals, suggesting cognitive dysfunction as an early sign of PD [8]. Among numerous pathophysiological mechanisms proposed for cognitive dysfunction in PD, dopamine deficiency is one of the most known hypotheses [9]. Deficiency in nigrostriatal dopamine was associated with executive dysfunction among patients with early-stage drug naïve PD [10]. Studies on healthy people have also proposed correlations between striatal DAT binding and cognitive dysfunction, including verbal function and executive/working memory impairments [11, 12]. However, studies on association between various domains of cognitive dysfunction and striatal DAT binding among healthy individuals are scarce and heterogeneous and remain to be elucidated.

Given the importance of understanding early biological alterations in PD, we hypothesized that inter-individual variation of striatal DAT binding among healthy elderly individuals could be a robust endophenotype for epigenomic studies. Therefore, we conducted an epigenome-wide association study (EWAS) and investigated the association between the whole blood DNA methylome profile and the inter-individual variation of striatal DAT binding measured by DaTscan among healthy individuals of the PPMI cohort. We also used functional omics approaches to prioritize the potential target genes in the identified loci. Given the importance of identifying the earliest clinical manifestations associated with the risk of PD, we also hypothesized that inter-individual variation of striatal DAT binding could manifest as mild cognitive impairment among healthy elderly individuals across specific domains. Thus, we explored the correlation of striatal DAT binding with several neuropsychological tests across five cognitive domains among healthy individuals. Our findings can bring a novel insight into the molecular and clinical aspects of early PD.

## 2. Methods

### 2.1. Study design and population

We used the PPMI cohort for our study. PPMI is a longitudinal, observational, and multi-center study that has collected various clinical, neuroimaging, and biological data from individuals across the PD spectrum. The consent forms have been obtained from all the participants. The details of this cohort can be found on its website (https://www.ppmi-info.org/).

Based on our research purpose, we included healthy individuals with no parkinsonism syndromes whose baseline DaTscan data were available. Then, we filtered subjects whose baseline cognitive tests or blood methylome data were available for subsequent analyses.

### 2.2. DaTscan

DaTscan was performed in PPMI imaging centers. The imaging protocol of the cohort is available at http://www.ppmi-info.org/study-design/research-documents-and-sops/. All imaging preprocessing steps were performed according to the PPMI protocol. Count densities were extracted from the caudate nucleus and putamen. The occipital cortex was also considered the reference region. The regional SBR was calculated as follows: (SBR)= (striatal region)/(occipital)−1 [13]. We focused only on mean, right, and left striatal DAT bindings. The mean striatal SBR was calculated by the following formula: Mean striatal SBR= (left caudate SBR+right caudate SBR+left putamen SBR+ right putamen SBR)/4.

### 2.3. Whole blood DNA methylation

Whole genome methylation of whole blood samples of the PPMI cohort was performed using the Illumina human methylation EPIC array at baseline visits. The methylation EPIC array covers >850,000 methylation sites across the whole genome (17 CpG sites per gene on average) at the single nucleotide resolution. We first downloaded the final normalized dataset of the PPMI cohort and separated the healthy individuals. Some quality control steps were performed using the Bioconductor package minfi to produce this dataset: Samples with a mean detection P-value >0.01 were excluded. Samples discordant between clinically reported sex and methylation-predicted sex were also excluded. Specific sentrix arrays were excluded for subjects who were processed more than once. Probes with a detection P value >0.01 in more than 20% of the samples were identified. Blood cell composition was determined using the minfi “estimateCellCounts” function, which assesses the relative proportion of CD8+ and CD4+ T cells, natural killer cells, B cells, monocytes, and granulocytes using the Houseman algorithm. Multidimensional scaling was also performed. Finally, functional normalization was performed on data using “preprocessFunnorm,” followed by removing 2769 failed probes. More details about the quality control steps can be found on the PPMI website. We conducted some additional quality control steps using the OSCA software (https://yanglab.westlake.edu.cn/software/osca/): We excluded CpG probes located on sex chromosomes and probes whose genomic positions were not available based on the “EPICv2.hg38.manifest” file provided by the Illumina company. Principal component analysis (PCA) was performed to detect outlier samples. We excluded samples whose first and second principal components were more than ±3 standard deviation from the means. Compared to a similar previous study [14], we adopted a more stringent quality control strategy: we excluded probes with a standard deviation smaller than 0.025, a missing ratio of more than 0.01, and a mean beta value of more than 0.85 or less than 0.15.

### 2.4. EWAS analysis to identify differentially methylated positions (DMPs)

For EWAS analysis, we first adjusted the beta values of CpG probes for age, sex, blood cell composition, and BMI for each sample. EWAS analysis was performed using the mixed linear model regression option (“moa”) of the OSCA software with mean, right, and left striatal DAT binding as the dependent variables and all methylation probes as independent variables. We used age, gender, and handedness as potential covariates affecting striatal DAT binding. Since our sample size was modest for EWAS analysis, we reported methylation probes with P-values of less than 1×10^−5^ as statistically suggestive, based on previous studies [15]. Since EWAS studies are prone to high inflation rates and bias, we first computed the inflation factor (λ) using the conventional approach. In the next step, we used the BACON package [16], specifically developed to control bias and inflation in EWAS studies, to obtain more accurate P-values. We finally reported the bias- and inflation-corrected test statistics computed by this package.

### 2.5. Differentially methylated region (DMR) analysis

We used the dmrff package [17] for DMR analysis. We used the default of the package. Briefly, this package identifies regions composed of methylation sites with EWAS P-values of less than 0.05 at a maximum 500 base-pair gap between CpG sites. Based on previous studies, we adopted a less stringent threshold of statistical significance and reported suggestive DMRs with P-values of less than 1×10^−4^ [15]. Additionally, we reported only regions containing at least three probes.

### 2.6. Positional and Functional mapping of DMPs

We first performed positional mapping of our significant DMPs using the “EPICv2.hg38.manifest” file provided by the Illumina website. We also searched for the overlap of our top DMPs with 90 PD GWAS loci previously reported in largest PD GWAS conducted by Nalls *et al*. [18]. We then searched for the overlap of significant DMPs with enhancers across B-cells, mononuclear phagocytes, and T-cells using the enhancer-gene map generated from 131 human cell types and tissues by the Activity By Contact (ABC) model [19]. To investigate the potential target genes of enhancer regions in the PD-related tissues, we used a single-nucleus multimodal dataset (snRNA-seq+snATAC-seq) obtained from the midbrain of 9 post-mortem healthy elderly individuals [20]. We first searched for the overlap of our EWAS DMPs identified in the pervious step with the chromatin-accessible regions. Due to the sparsity nature of single-cell data, we then used the Spearman correlation test to investigate the potential correlation between the expression of nearby genes at the EWAS top DMPs (±200kb) with the chromatin accessibilities across all cells of each sample. We adopted a correlation coefficient of more than 0.30 as a significant correlation.

### 2.7. Cognitive phenotypes

We extracted 12 test scores across five cognitive domains from healthy individuals of the PPMI cohort. These 12 clinical phenotypes were categorized as follows: Executive/working memory domain (letter number sequence test, semantic fluency animal sub score test, and trail making test B), language/executive domain (lexical fluency test and modified Boston naming test), attention/processing speed domain (symbol digit modalities test and trail-making test A), visuospatial processing domain (Benton judgment of line orientation test and clock drawing test), and memory domain (Hopkins verbal learning (HVL) immediate recall test, HVL delayed recall test, and HVL discrimination recognition test). Since data from healthy individuals of the PPMI cohort have been collected in various phases, the results of 5 tests (lexical fluency test, trail-making test A and B, modified Boston naming test, and clock drawing test) were not assessed among the healthy individuals of the original cohort which resulted in smaller sample sizes relative to other tests. Then, we transformed the scores of 12 cognitive tests into Z-scores (mean 0 and standard deviation=1) by dividing the difference of scores from the mean to the standard deviation of scores.

### 2.8. Investigating association of cognitive phenotypes with striatal DAT binding

To investigate subtle impairments of which cognitive domain may reflect striatal DAT binding variation among healthy individuals, we used multiple separate linear regression models for each of the 12 neuropsychological tests, with the Z-score of neuropsychological tests as the dependent variables and the mean, left, and right striatal DAT binding as the independent variables. The results were adjusted for age, gender, and education. Given the exploratory nature of the analysis, we deemed a P-value of less than 0.05 statistically significant. All the analyses were performed by the R version 4.3.

## 3. Results

### 3.1. EWAS results

The whole blood DNA methylome data of 865,859 CpG probes among 420 participants at their baseline visit were available in the PPMI cohort dataset. Among these subjects, 99 were healthy individuals. We included 96 healthy individuals whose DaTscan and BMI data were available. We then performed additional quality controls on these individuals. First, we excluded 19,663 probes due to their location on the sex chromosomes or missing genomic positions. No outlier was identified based on the top 2 principal components of the methylation profile. Then, 369,632 probes with standard deviations of less than 0.025 were excluded. 97,406 probes with mean beta values of less than 0.15 or more than 0.85 were also excluded. The missing ratio of no probes was more than 0.01. Finally, we adjusted the beta values of CpG probes for age, sex, blood cell compositions, and BMI for each sample. The mean age of the subjects was 61.75 (±11.04).

Twenty-five subjects (26.04%) were female, and 71 were male. The mean of mean, right, and left striatal DAT bindings were 2.48 (±0.51), 4.94 (±1.03), and 4.98 (±1.05), respectively. Finally, we performed EWAS using 379,158 CpG probes from 96 healthy individuals using age, gender, and handedness as covariates. We did not observe significant deflation or inflation in our three analyses (λ_bacon_=1.01, 1.04, and 1.05). We identified seven DMPs associated with striatal DAT binding with P-values of less than 1×10^−5^. The complete summary statistics of top seven DMPs are shown in Table 1. The Manhattan and Q-Q plots of our EWAS are depicted in Figure 1. Among these seven DMPs, three (cg18316254, cg08118333, and cg14458991) were located in close vicinity of three previously reported GWAS hits in the largest PD GWAS to date. The genomic locations of two previously reported PD GWAS hits and our EWAS results are also shown in Figure 2.

**Table 1.**
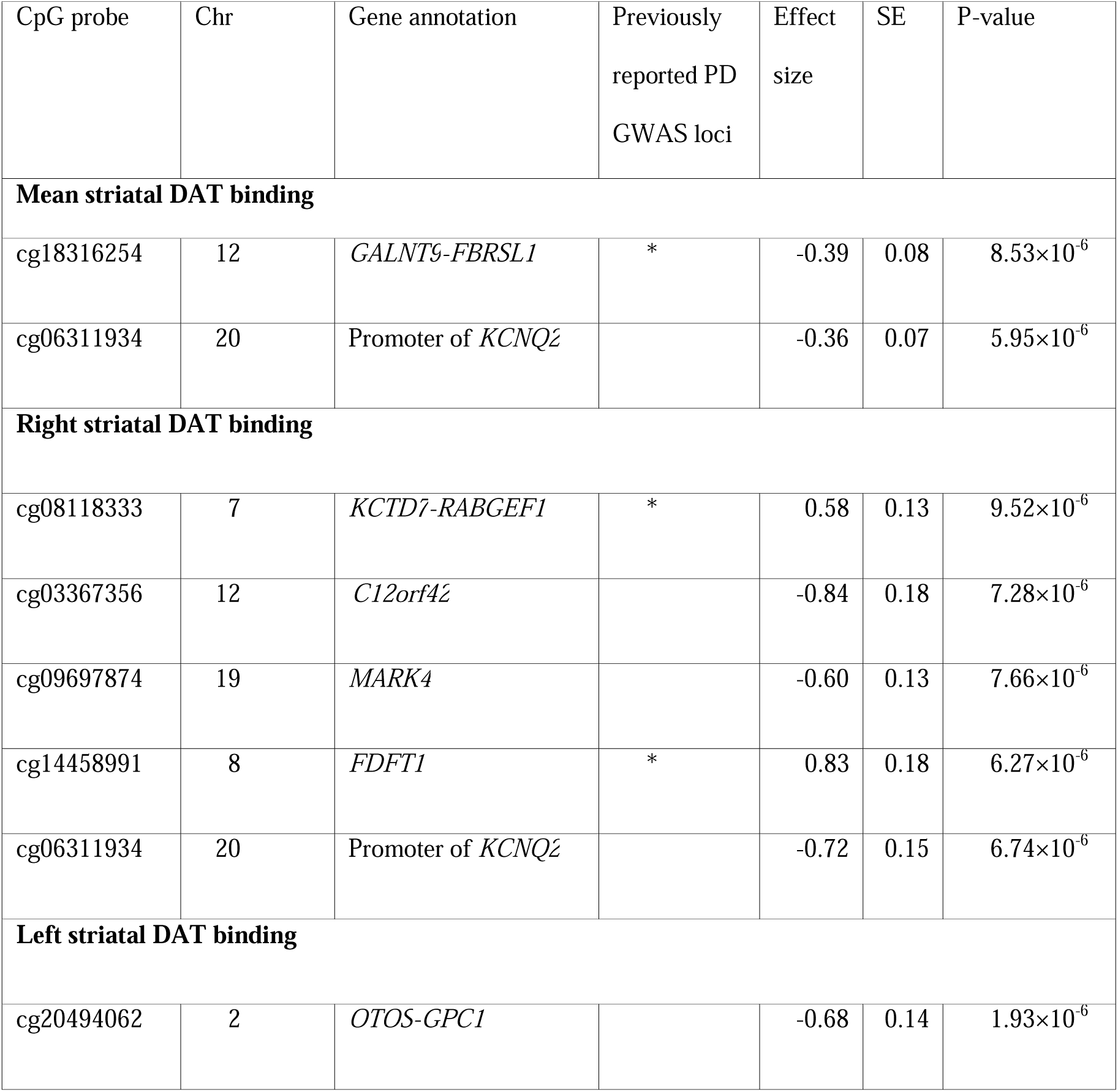
EWAS Summary statistics of top 7 DMPs associated with striatal DAT binding.

| CpG probe | Chr | Gene annotation | Previously reported PD GWAS loci | Effect size | SE | P-value |
| --- | --- | --- | --- | --- | --- | --- |
| **Mean striatal DAT binding** |  |  |  |  |  |  |
| cg18316254 | 12 | *GALNT9-FBRSL1* | * | -0.39 | 0.08 | $8.53 \times 10^{-6}$ |
| cg06311934 | 20 | Promoter of *KCNQ2* | | -0.36 | 0.07 | $5.95 \times 10^{-6}$ |
| **Right striatal DAT binding** |  |  |  |  |  |  |
| cg08118333 | 7 | *KCTD7-RABGEF1* | * | 0.58 | 0.13 | $9.52 \times 10^{-6}$ |
| cg03367356 | 12 | *C12orf42* | | -0.84 | 0.18 | $7.28 \times 10^{-6}$ |
| cg09697874 | 19 | *MARK4* | | -0.60 | 0.13 | $7.66 \times 10^{-6}$ |
| cg14458991 | 8 | *FDFT1* | * | 0.83 | 0.18 | $6.27 \times 10^{-6}$ |
| cg06311934 | 20 | Promoter of *KCNQ2* | | -0.72 | 0.15 | $6.74 \times 10^{-6}$ |
| **Left striatal DAT binding** |  |  |  |  |  |  |
| cg20494062 | 2 | *OTOS-GPC1* | | -0.68 | 0.14 | $1.93 \times 10^{-6}$ |

**Figure 1.**
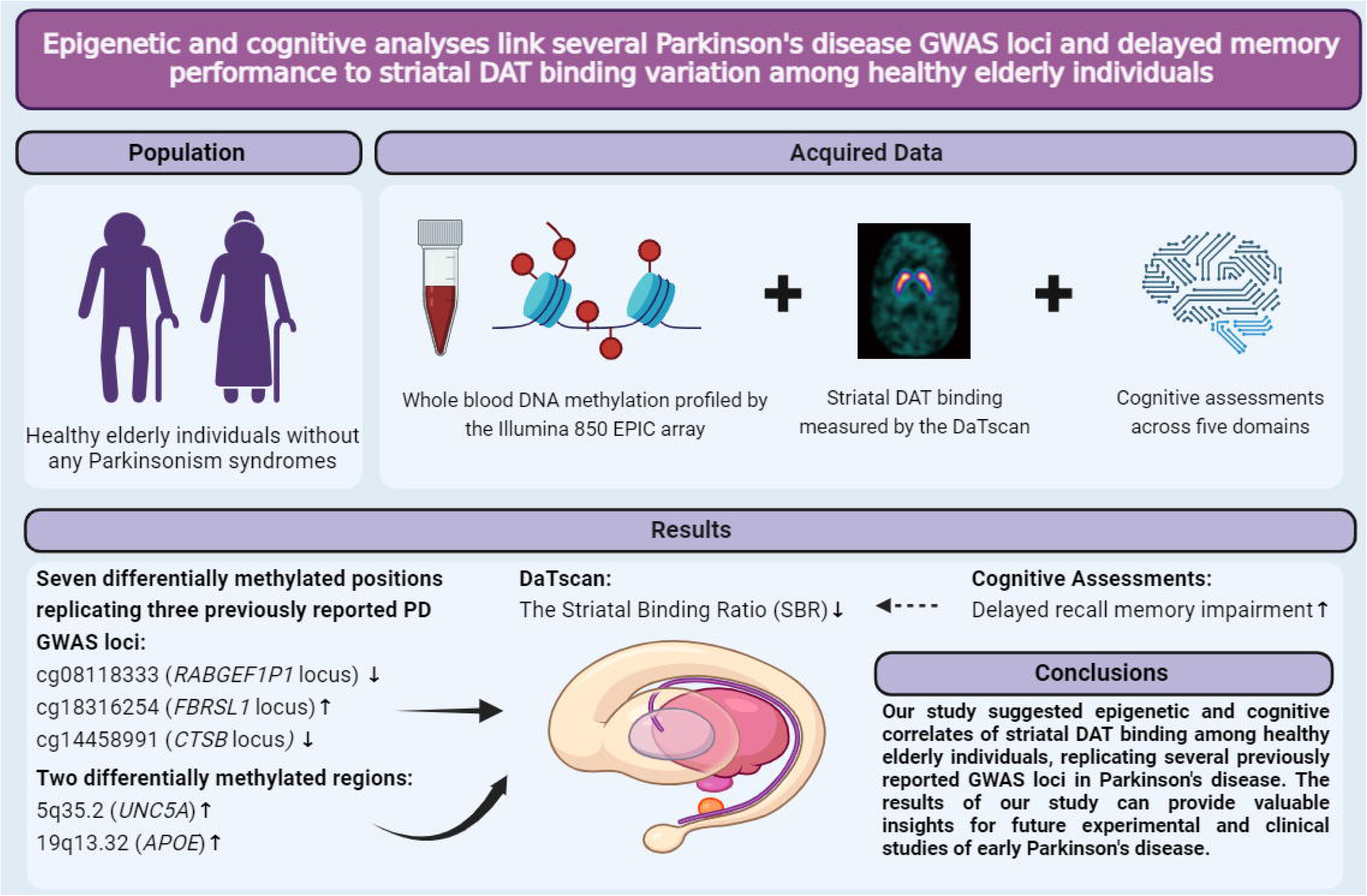
The figures depict the Manhattan and Q-Q plots of mean, right, and left striatal DAT binding EWAS among 96 healthy individuals. Each figure has two panels: a) This panel shows the Manhattan plot with the positional gene mapping of the top DMPs. The blue line shows the suggestive threshold (P-value<10^−5^) of our EWAS. Loci previously reported as top PD GWAS hits are highlighted in green. b) This panel illustrates the Q-Q plot of the P-values of our EWAS results with the bias- and inflation-corrected lambda of our analysis computed by the BACON package.

**Figure 2.**
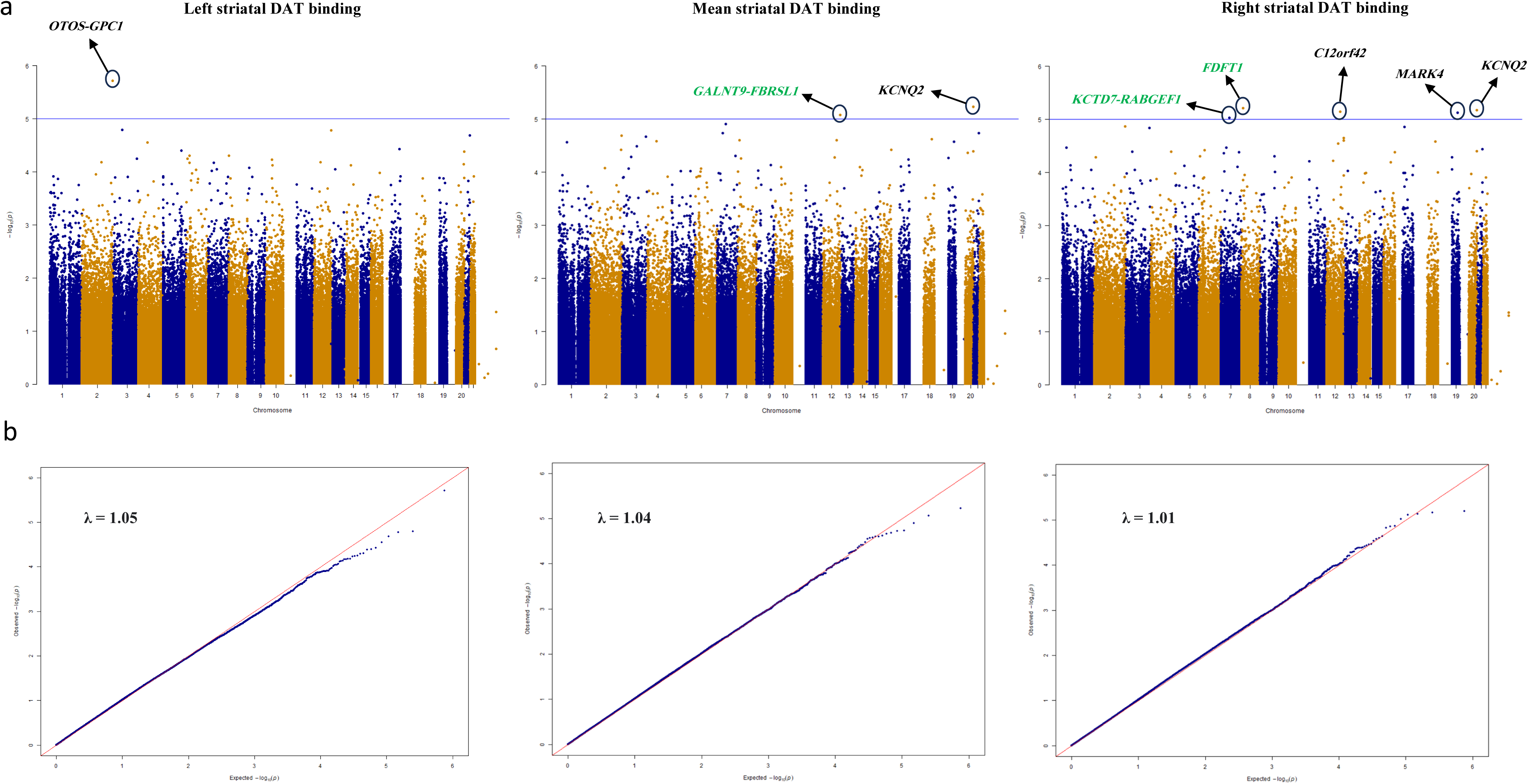
The figure shows two top significant methylation probes of our EWAS on striatal DAT binding and their positional position and vicinity to two previously reported GWAS hits of the largest PD GWAS to date: a) This panel zooms into one of the top GWAS hits of the Nalls et al. study in the 7q11.21 locus and its linkage disequilibrium (LD) structure. The top SNP rs76949143 in this locus lies between the *TPST1* and *KCTD7* genes. It also shows the location of our top EWAS result (cg08118333 methylation probe) between the *KCTD7* and *RABGEF1* genes. The lower part shows the direct linear correlation of cg08118333 methylation with right striatal DAT binding among 96 healthy individuals obtained from our EWAS analysis. b) This panel zooms into one of the other top GWAS hits of the Nalls *et al*. study in the 12q24.33 locus and its LD structure. The top SNP rs11610045 in this locus is located between the *GALNT9* and *FBRSL1* genes. It also shows the location of our top EWAS result (cg18316254 methylation probe) between the *GALNT9* and *FBRSL1* genes. The lower part shows the inverse linear correlation of cg18316254 methylation with mean striatal DAT binding among 96 healthy individuals obtained from our EWAS analysis.

### 3.2. DMR analysis results

In all three analyses, hypermethylation of two regions were suggestively (1×10^−4^) associated with lower striatal DAT binding: A region on 5q35.2 in UNC5A gene (min P-value=9.98×10^−6^ associated with right striatal DAT binding) containing three CpG probes and a region on 19q13.32 in CEACAM20 gene (APOE locus) (min P-value= 1.78×10^−5^ associated with mean striatal DAT binding) containing three CpG probes.

### 3.3. Functional annotation of DMPs

Among our top DMPs, only one DMP (cg14458991) was located in enhancer regions predicted by the ABC method across blood cells. Three target genes with the highest ABC score in B-cells were identified for this region: The *FDFT1*, *CTSB*, and *NEIL2* genes. The list of genes in the ABC model and the genomic location of the enhancers for this DMP are shown in Supplementary Figure 1. We then found that this DMP overlapped with chromatin-accessible regions in the single-nucleus ATAC-seq of the midbrain regions of 9 post-mortem healthy elderly individuals. Multiple correlational analyses between chromatin accessibility and gene expression of four nearby genes in this locus (*FDFT1*, *CTSB*, *NEIL2*, and *GATA4*) prioritized the *FDFT1* gene as the potential target gene of the enhancer. The *FDFT1* gene was the top gene across all 9 samples, and its expression was significantly correlated with the ATAC peak region (max correlation coefficient=0.32). The complete results of our functional analyses, EWAS result of this DMP, and genomic position of this DMP in the context of the GWAS hit in this locus are shown in Figure 3.

**Figure 3.**
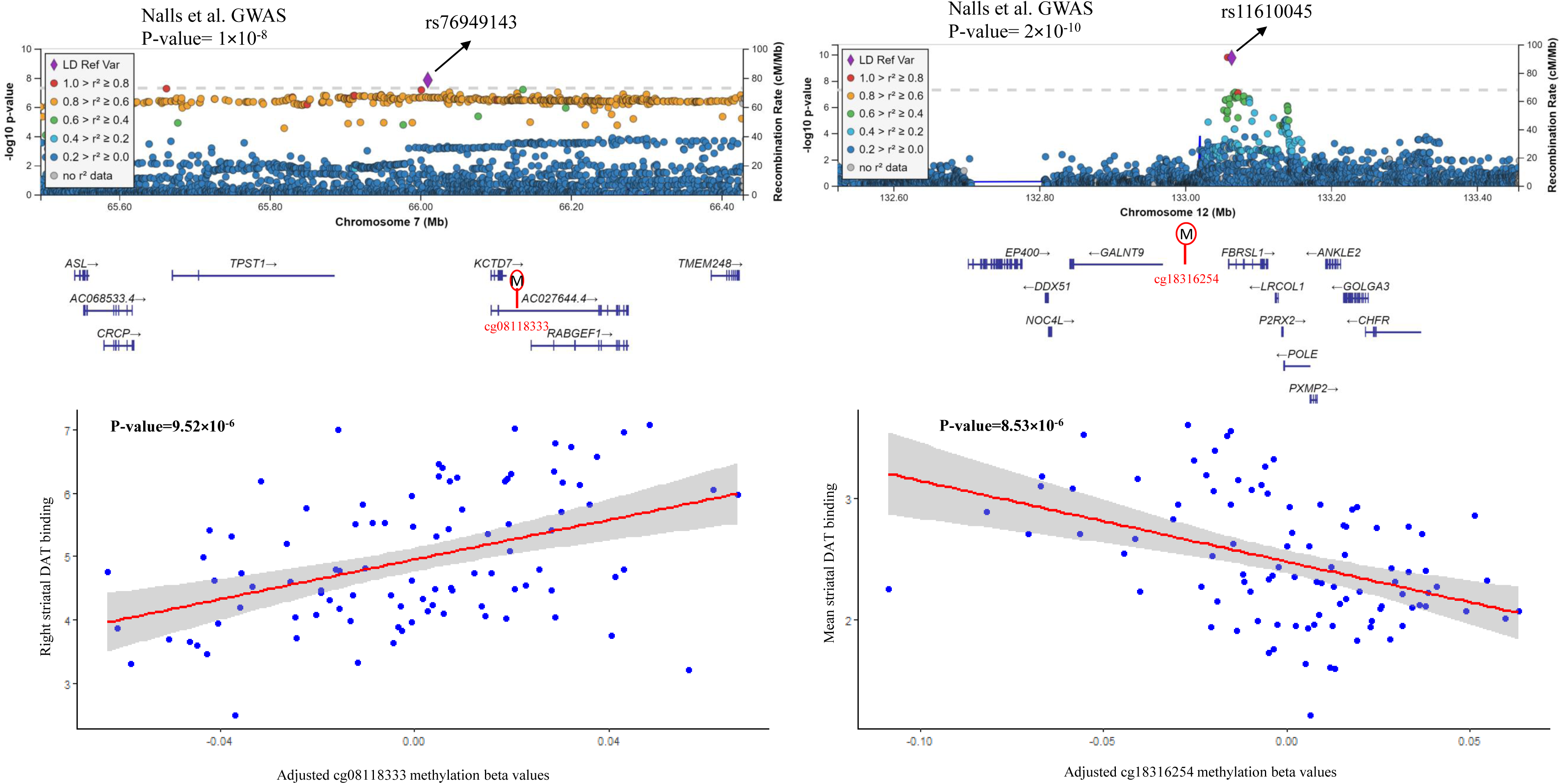
The upper part of the figure shows the genomic position of one of the suggestive DMPs associated with right striatal DAT binding and its vicinity to the *CTSB* locus previously reported in the largest PD GWAS. The lower parts illustrate the overlap of this DMP with the chromatin-accessible region of the midbrain of nine healthy elderly individuals. It also shows our enhancer-gene mapping strategy using a midbrain single-nucleus multimodal dataset, which prioritized the *FDFT1* gene as the potential candidate gene in this PD GWAS locus. a) This panel zooms into the *CTSB* GWAS locus, previously reported in the largest PD GWAS conducted by Nalls *et al*., and its LD structure and P-value. The top SNP rs1293298 in this locus is located in the *CTSB* gene. It also shows the location of our top EWAS DMP (cg14458991 methylation probe) in the *FDFT1* gene next to the *CTSB* gene and a previously reported DMP in PD EWAS (cg17155499) in the *NEIL2* gene. The lower part shows the overlap of the cg14458991 with the ATAC peaks obtained from the single-nucleus multimodal dataset of midbrain of 9 post-mortem healthy elderly individuals. b) This panel shows the direct linear correlation of the cg14458991 methylation probe with right striatal DAT binding among 96 healthy individuals obtained from our EWAS analysis. c) The lower part of the panel shows the heatmap result of our enhancer-gene mapping analysis. The rows are four potential genes in this locus. The columns show nine multimodal samples obtained from the midbrain of post-mortem healthy elderly individuals. Each cell’s colors depict the degree to which ATAC peak regions correlate with gene expression. The upper part shows the scatter plot of the correlation between the expression of the top gene, *FDFT1*, and the chromatin accessibility of that region.

### 3.4. Results of cognitive function analysis

The results of seven neuropsychological tests from 252 healthy individuals with DaTscan data were extracted. We also extracted the results of five neuropsychological tests from 62 healthy individuals from the other phase of the cohort whose DaTscan data were also available. The association of mean, right, and left striatal DAT binding with these neuropsychological tests were separately investigated. A few missing data were found in a few tests and excluded from the analysis. The mean age of 252 subjects was 61.53 (±11.64). Ninety-six (38.09%) were female, and 156 were male. The mean of mean, right, and left striatal DAT binding were 2.62 (±0.55), 5.22 (±1.10), and 5.26 (±1.12), respectively. The mean of educational years was 16.11 (±3.08). Among the 62 healthy individuals, the mean age was 64.22 (±12.39). Thirty-four were male (54.83%), and 28 were female. The mean of the mean, right, and left striatal DTA bindings were 2.77 (±0.49), 5.50 (±1.01), and 5.59 (±1.01), respectively. The mean of the educational years was 16.37 (±3.59). The number of subjects for each analysis is shown in Table 1. Only two of 12 neuropsychological tests were significantly correlated with mean and right striatal DAT bindings: HVLT delayed recall test (spearman correlation coefficient=0.22, P-value= 4×10^−4^) and HVLT discrimination recognition test (spearman correlation coefficient=0.20, P-value=10^−3^). Among these two tests, only the HVLT delayed recall test remained statistically significant after adjusting for potential confounders, including age, gender, and education (P-value=0.04). The complete results of various cognitive tests associations with the mean striatal DAT binding are shown in Table 2.

**Table 2.** Association of striatal DAT binding with a battery of neuropsychological tests across 5 cognitive domains.

| Cognitive domains | Number of subjects | Mean score (±SD) | Age |  | Education |  | Gender |  | Striatal DAT binding |  |
| --- | --- | --- | --- | --- | --- | --- | --- | --- | --- | --- |
| Executive-working memory |  |  | P-value | Effect size (SE) | P-value | Effect size (SE) | P-value | Effect size (SE) | P-value | Effect size (SE) |
| Letter number sequencing | 251 | 10.86 (±2.71) | **3.00×10<sup>-6</sup>** | -0.30 (0.07) | **0.004** | 0.18 (0.07) | 0.68 | 0.02 (0.06) | 0.72 | -0.02 (0.05) |
| Semantic fluency animal sub score | 251 | 22.16 (±5.32) | 0.37 | -0.05 (0.06) | 0.09 | 0.10 (0.06) | 0.25 | -0.07 (0.06) | 0.41 | 0.05 (0.06) |
| Trail making test B | 61 | 74.42 (±42.26) | 0.09 | 0.22 (0.22) | 0.96 | -0.005 (0.08) | 0.46 | -0.10 (0.14) | 0.93 | -0.01 (0.14) |
| Language-executive |  |  |  |  |  |  |  |  |  |  |
| Lexical fluency | 61 | 44.73 (±12.71) | 0.71 | -0.04 (0.10) | **0.02** | 0.30 (0.10) | 0.56 | 0.08 (0.14) | 0.72 | 0.05 (0.14) |
| Modified | 61 | 55.75 | 0.64 | 0.05 | 0.05 | 0.25 | 0.19 | 0.18 | 0.11 | 0.22 |
| Boston<br>naming test |  | (±7.85) |  | (0.10) |  | (0.10) |  | (0.14) |  | (0.14) |
| **Attention-processing speed** |  |  |  |  |  |  |  |  |  |  |
| Symbol<br>digit<br>modalities<br>test | 251 | 47.31<br>(±10.37) | **5.68**×10<sup>-19</sup> | -0.53 (0.06) | **0.006** | 0.15<br>(0.03) | 0.14 | -0.08<br>(0.05) | 0.86 | 0.01<br>(0.10) |
| Trail<br>making test<br>A | 61 | 32.98<br>(±11.99) | **0.02** | 0.30<br>(0.15) | 0.65 | 0.05<br>(0.16) | 0.46 | -0.10<br>(0.14) | 0.97 | 0.005<br>(0.14) |
| **Visuospatial processing** |  |  |  |  |  |  |  |  |  |  |
| Clock<br>drawing test | 62 | 6.38<br>(±1.23) | 0.31 | -0.13<br>(0.13) | 0.33 | -0.13<br>(0.13) | 0.51 | 0.09<br>(0.14) | 0.44 | 0.11<br>(0.14) |
| Benton<br>judgment of<br>line<br>orientation<br>test | 250 | 13.14<br>(±2.04) | **0.04** | -0.12 (0.06) | **0.001** | 0.20<br>(0.06) | **5.90**×10<sup>-5</sup> | 0.25<br>(0.05) | 0.44 | -0.04<br>(0.05) |
| **Memory** |  |  |  |  |  |  |  |  |  |  |
| HVLT<br>immediate<br>recall | 251 | 26.01<br>(±4.58) | **2.97**×10<sup>-4</sup> | -0.23 (0.05) | 0.12 | 0.09<br>(0.06) | 0.05 | -0.12<br>(0.06) | 0.91 | 0.007<br>(0.07) |
| HVLT<br>delayed<br>recall | 251 | 9.24<br>(±2.34) | **0.001** | -0.20 (0.10) | **0.04** | 0.12<br>(0.06) | **0.04** | -0.12<br>(0.06) | **0.04** | 0.13<br>(0.06) |
| HVLT | 251 | 10.14 | 0.61 | -0.03 (0.06) | 0.83 | -0.01 | 0.27 | -0.07 | 0.05 | 0.12 |
| discriminati<br>on<br>recognition |  | (±2.64) |  |  |  | (0.05) |  | (0.06) |  | (0.06) |

## 4. Discussion

To the best of our knowledge, this is the first study investigating the epigenetic signatures of striatal DAT binding among healthy individuals. Using whole blood methylome data from healthy individuals, we found that the altered methylations of seven loci, including three previously reported PD GWAS loci, were significantly associated with lower striatal DAT binding. Using a single-nucleus multimodal dataset obtained from the midbrain of post-mortem healthy elderly individuals, we prioritized the *FDFT1* gene as the candidate gene in the previously reported *CTSB* GWAS locus. Finally, multiple investigative linear regression analyses unveiled that reduced striatal DAT binding was markedly correlated with subtle impairments in delayed memory function among healthy individuals.

We found several DMPs lying at loci potentially related to PD pathology. For instance, we found that hypermethylation at the promoter of the Potassium Voltage-Gated Channel Subfamily Q member 2 (*KCNQ2*) gene correlated with reduced striatal DAT binding among healthy individuals. Growing evidence has shown that potassium channels regulate the firing of dopaminergic neurons in the striatum. The expressions of two members of the *KCNQ* family (*KCNQ2* and *KCNQ4*) are mostly confined to the substantia nigra and ventral tegmental area of the midbrain based on immunohistochemistry studies [21]. Interestingly, modulation of the *KCNQ2* has been proposed as a potential therapeutic target for PD [22]. Regarding the cg20494062 located between the *OTOS* and *GPC1* genes, a previous study has shown that the rs4417745 between *OTOS* and *GPC1* genes was associated with frontotemporal dementia disease [23]. However, due to the intergenic nature of most of these DMPs, future functional studies are needed to unravel the exact target genes in these loci.

Interestingly, we found that among our top seven suggestive DMPs, three were located in close vicinity of three GWAS hits previously reported in the largest PD GWAS conducted by Nalls *et al*. [18]. For instance, regarding the 08118333 probe, both *KCTD7* and *RABGEF1* genes can be involved in PD. By regulating K^+^ conductance through K^+^ channels, the *KCTD7* gene is involved in neuronal stimulation and activation [24]. Using mammalian cultured cells, Yamano and colleagues have shown that the RABGEF1 protein directly interacts with Parkin, one of the most famous mutated genes implicated in familial PD, in mitophagy-related pathways [25]. Our study could replicate these two known PD GWAS loci at the methylome level, this time in association with striatal DAT binding among healthy individuals. These findings can shed light on the biological pathways involved in early PD.

We found two suggestive DMRs associated with all striatal DAT binding measures. The first one was located on 19q13.31 in the *CEACAM20* gene in the *APOE* region, which was reported previously in Alzheimer’s disease (AD) GWAS [26]. The other region was located on 5q35.2 in the *UNC5A* gene. The UNC5A gene encodes one of the netrin receptors and is involved in axonal guidance and nervous system development. Apoptotic netrin UNC5 signaling has been implicated in AD and PD pathology [27].

Another DMP identified in our EWAS, cg14458991, is located next to the previously reported *CTSB* gene in the largest PD GWAS. Several interesting genes exist in this region, such as the *FDFT1*, *CTSB*, *NEIL2,* and *GATA4*. A recent study has comprehensively examined this region and shown experimentally that the *CTSB* gene is the main driver of PD in this locus by inhibiting the formation of alpha-synuclein fibrils in the dopaminergic neurons [28]. Expression quantitative trait loci (eQTL) studies have shown that the GWAS hit (rs1293298) affects the expression of the *FDFT1*, *CTSB*, and *NEIL2* genes in blood tissue (https://fivex.sph.umich.edu/). Using the ABC model, we found that across B-cells, the *FDFT1*, *CTSB*, and *NEIL2* were the potential target genes of this potential enhancer region. Our enhancer-gene mapping strategy prioritized the *FDFT1* gene as the potential target gene of this region in the midbrain. The *FDFT1* gene product is the first specific enzyme in cholesterol synthesis, one of the pathways linked to PD and alpha-synuclein aggregation [29–31]. Interestingly, a blood-based methylome-wide association study with 1132 patients with PD and 999 control subjects indicated that differential methylation of the cg17155499 locus in the *NEIL2* gene, a neighbor of the *FDFT1* gene, is associated with PD [32]. Future experimental studies are needed to unravel the exact driver of PD in this region. However, the association of altered methylation of this previously reported locus in PD with striatal DAT binding, even among healthy elderly individuals, can be insightful for understanding early pathological alterations in PD.

Assessing clinical cognitive functions, we found that striatal DAT binding is markedly lower in healthy individuals with subtle impairments in RAVLT delayed recall memory, irrespective of the effect of age, gender, and education. It was reported that impairments of memory and executive function domains are the prominent cognitive deficits in patients with newly diagnosed PD [33]. A few studies have shown a connection between striatum presynaptic dopamine function and memory performance [34]. In line with our findings, Chen *et al.* showed that delayed verbal memory and working memory explain most of the variation of the D2/D3 receptor densities in the striatum of 62 healthy individuals [35]. However, due to the scarcity of clinical imaging studies on non-PD individuals (healthy and prodromal elderly individuals), more studies are needed to investigate impairments in which the cognitive domain manifests earlier during the PD course.

Our study has some limitations: Our sample size was modest. However, since our primary goal was to generate novel hypotheses on the pathophysiological mechanisms involved in early PD, our population can be unique, considering the high costs of obtaining DaTscan and blood methylome profiles from healthy individuals. However, more sample sizes are needed in future studies to confirm or rule out our findings. Although we did not validate the result of blood EWAS in other healthy cohorts, replicating three PD GWAS loci in our EWAS may indicate the robustness of our findings. We hope that the suggestive results of our study, at molecular and clinical levels, bring novel insights for uncovering the pathological mechanisms underlying the earliest stages of PD in future studies.

In conclusion, our blood EWAS revealed that altered methylations of seven distinct CpG sites were associated with lower striatal DAT binding among healthy individuals, highlighting the replication of three previously reported PD GWAS loci at the methylome level, even among healthy individuals. Using a midbrain single-nucleus multimodal dataset, we also prioritized the *FDFT1* gene as the potential target gene in the *CTSB* GWAS locus. We also showed that lower striatal DAT binding can manifest as subtle impairments in delayed memory function among healthy individuals.

## Declarations

## Acknowledgment

None.

## Ethics approval and consent to participate

Not applicable.

## Consent for publication

Not applicable.

## Availability of data and materials

We used data from the PPMI cohort for this study. The details of PPMI cohort cohort can be found on its website (https://www.ppmi-info.org/). By sending our research proposal form, we had permission to access the database of the PPMI cohort via the LONI database (https://ida.loni.usc.edu/).

## Authors’ contribution

AY (conceptualization, data analysis, and writing and editing the draft), HS (writing and editing the draft), MJ (writing the draft), and MA (editing the draft). All authors studied the final version of the article and approved this submission.

---

**Supplementary Figure 1.**
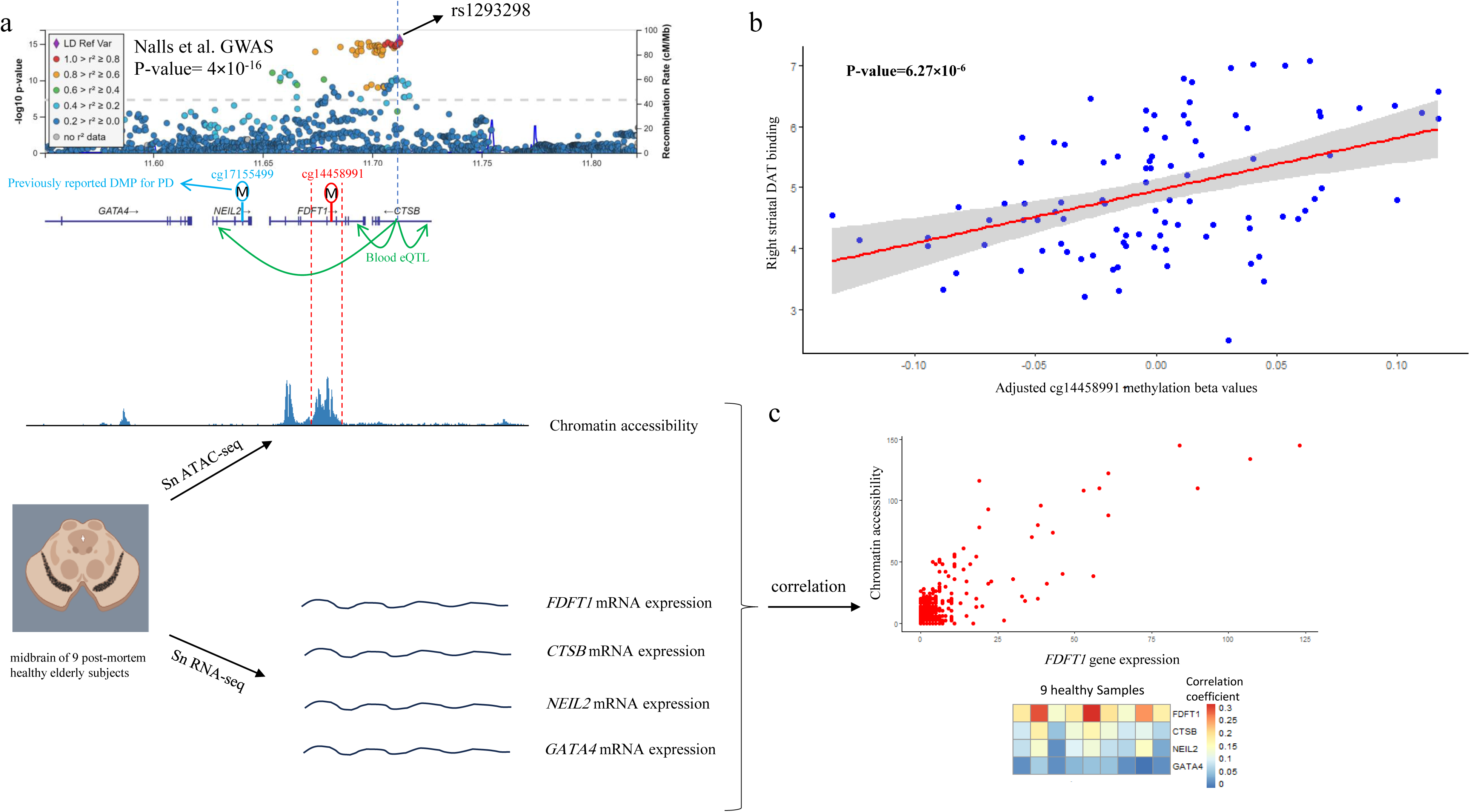
The figure shows the genomic position of one of the DMPs (cg14458991) and the enhancers in that region predicted by the ABC model across blood cells: a) This panel illustrates the overlap of enhancer regions, predicted by the ABC model across three blood cells (mononuclear phagocytes, B-cells, and T-cells), with the cg14458991 methylation probe. b) This panel shows the target genes of cg14458991 predicted by the ABC model based on the overlap of this DMP with enhancers across B and T cells. b) This panel shows the direct linear correlation of the cg14458991 methylation probe with right striatal DAT binding among 96 healthy individuals obtained from our EWAS analysis. c) The lower part of the panel shows the heatmap result of our enhancer-gene mapping analysis. The rows are four potential genes in this locus. The columns show nine multimodal samples obtained from the midbrain of post-mortem healthy elderly individuals. Each cell’s colors depict the degree to which ATAC peak regions correlate with gene expression. The upper part shows the scatter plot of the correlation between the expression of the top gene, *FDFT1*, and the chromatin accessibility of that region.

