## Supplementary figures and images for "Epigenetic and cognitive signatures of striatal DAT binding among healthy elderly individuals"

### Supplementary Figure 1

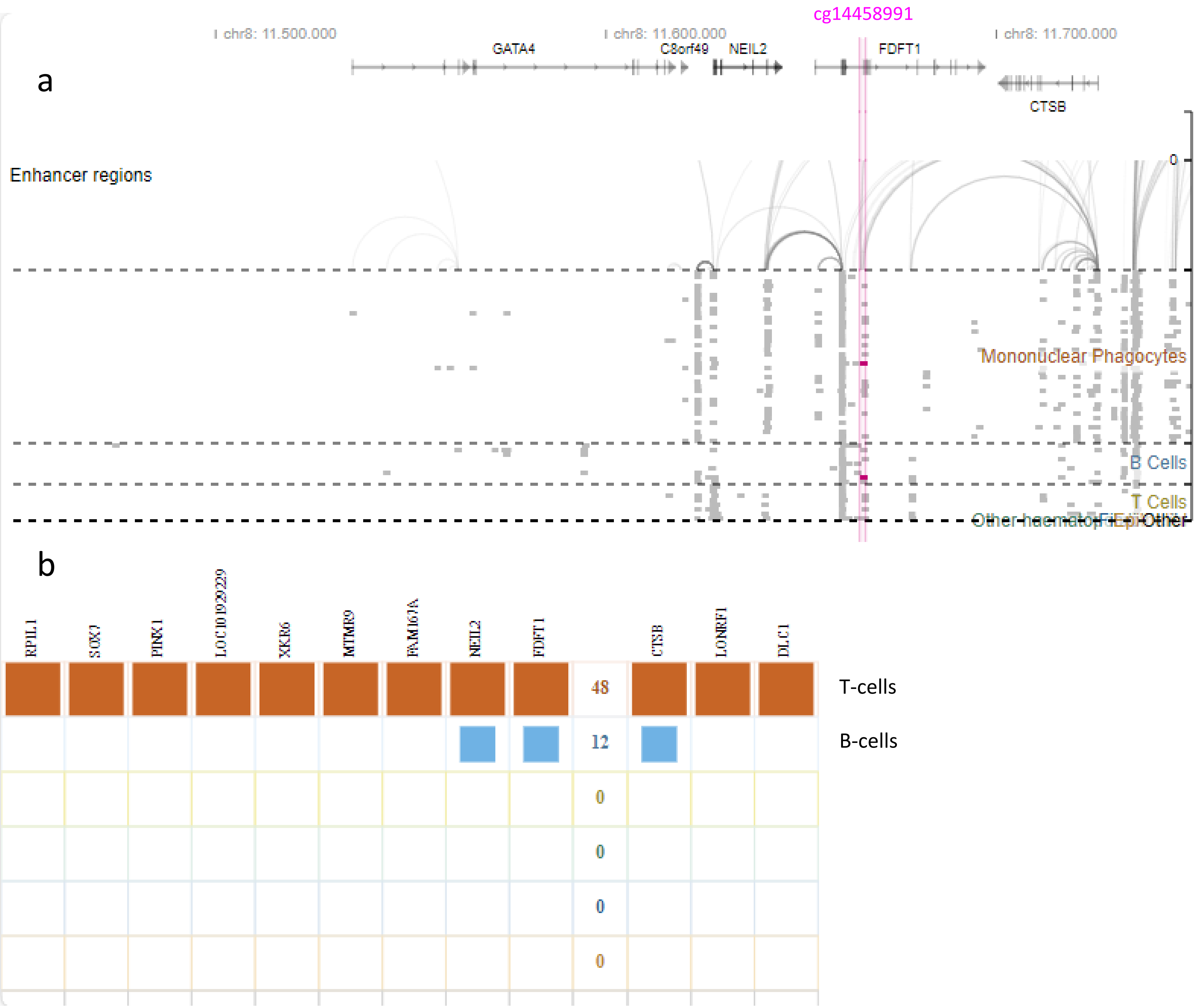
